# The Healthy Human Global Project - Hong Kong : a community-based cross-sectional study of a healthy Asian population

**DOI:** 10.1101/2024.09.12.24313504

**Authors:** Rex L Hung, Wilson W Ng, Peter K C Chung, Ashley B S Li, Theodora H Y Luk, Rosanna Fong, Anna Lee, Raina Mok, Hebronson Yung, Vera Tse, Raven Cheng, Amandine Schneider, Rachel L Telford, Florian Dubois, Elsa Bourayou, Milieu Interieur Consortium, Rafael De Andrade Moral, Jean Marc Doisne, Milena Hasan, Gabriel M Leung, Michael Y Ni, Michael Tse, Malik Peiris, James Di Santo, Roberto Bruzzone, Vincent Rouilly, Darragh Duffy

**Author notes:** equal contributions (Rex L Hung, Wilson W Ng). equal contributions (Peter K C Chung, Ashley B S Li, Theodora H Y Luk).

## Abstract

Immune responses are highly variable from one individual to another, with this variability determined by factors such as age, sex, genetics and environment, which also affect physical and mental well-being. Systems immunology studies have been applied to human cohort studies to better understand these effects, but most are performed in Western populations of mostly European ancestry, neglecting the vast majority of human global diversity. To overcome this limitation, we established the Healthy Human Global Project – Hong Kong (HHGP-HK) cohort to allow the study of immune variability in a healthy Asian population. Taking inspiration from the established French Milieu Interieur study we adapted inclusion and exclusion criteria to identify healthy donors, and to facilitate cross comparison between the two cohorts we collected similar demographic, medical and lifestyle information. To study the effects of age and sex, donors were stratified for both ranging from 20-79 years. We report here significant age and sex effects on multiple clinical laboratory measures, many of which were consistently observed in the Milieu Interieur cohort. C-reactive protein and liver enzymes were two exceptions perhaps reflecting different environmental effects between the two cohorts. We applied biological aging, physiological, and mental health scores to our cohort which highlights the need to develop and adapt such scores for application to populations of non-European descent. We also included wearable fitness trackers from which we observed significant age and sex differences for sleep, physical activity, and heart rate variability. This unique cohort will enable the better understanding of factors that determine immune variability within Asian populations.

## 1. Introduction

Immune responses are highly variable between individuals as a result of diverse genetic and environmental factors, as well as age and sex differences^1^. In recent years human cohort studies have been established to provide a better understanding of such factors, and specifically which immune responses they influence. Studies such as Milieu Interieur^2^, the Human Functional Genomics (FG500) project^3^, the SardiNIA study population cohort^4^, and the Stanford 1000 Immunomes Project^5^ have identified and quantified how factors such as age, sex, genetics, environmental and lifestyle factors influence variable immune responses. Other larger population based cohort studies such as the UK biobank^6^, FinnGen^7^, Lifelines^8^, and the All of Us Research Program^9^, have also enabled further understanding of how genetic factors impact clinical and health care related outcomes in the general population. However, the vast majority of such studies have been conducted in Europe or the US with strong inherent biases for European ancestry and western environmental effects. To capture the full extent of human diversity requires the study of human cohorts in other populations and geographical locations ^10^.

Host genetics has a substantial influence on immune variation, possibly as a result of selective evolutionary pressures driven by interactions with microbes^11^. Single nucleotide polymorphisms (SNP) have been estimated to account for between 0-79% of variability observed in specific immune responses^1^. However many of these investigations have either been conducted in small twin studies^12^, or populations of limited genetic diversity^3,4,13^. Some trans-ethnic analyses have been performed, with one study in South Asians identifying 71 novel associations with lymphocyte counts, including a missense variant in IL7, that were previously overlooked in studies of more homogenous European ancestry populations^14^. As well as missing such potential population specific genetic associations^15^, these studies are also limited for the understanding of gene-by-environment interactions. Furthermore biological scores of healthy aging are increasingly proposed as biomarker tools to monitor health and disease development^16^. However due to the Western bias in most data sets that are used to generate these biological scores they may not be relevant for applications in other populations.

To address some of these limitations we established the Healthy Human Global Project – Hong Kong (HHGP-HK) to provide a resource for studying healthy immune responses, and their determinants in an Asian population and environment. Taking advantage of previous work conducted by the Milieu Interieur consortium in France^2^, we adapted inclusion and exclusion criteria to this specific setting to recruit a cohort of healthy adult donors balanced for sex (half women, half men) and age (from the ages 20-79 years old). We also benefitted by involving an existing longitudinal local cohort in participant recruitment, the FAMILY Cohort^17^ which was established in 2009 to better understand the health and wellbeing of Hong Kong residents. Herein we describe the study design and recruitment of the cohort, report effects of age and sex on blood biological measures, and directly compare different biological measures and biomarker scores with a French healthy cohort (Milieu Interieur). Our findings emphasize the need and value in extending such cohort-based studies to less studied non-Western based populations to support the development of more tailored treatments and therapies in diverse populations.

## 2. Materials & methods

### 2.1 Clinical study design

The HHGP-HK cohort study is composed of 1,025 healthy Chinese volunteers recruited in Hong Kong stratified according to sex (512 female subjects and 513 male subjects); and age (6 decades of age: [20–29], [30–39], [40–49], [50–59], [60–69] and [70–79] years, with roughly 170 subjects per stratum). Donors self-declared Han ethnicity (the largest Chinese ethnic group in Hong Kong and mainland China) for 3 generations, with no other direct family member already involved, to control for genetic heterogeneity and increase statistical power to identify common variant associations. Approximately one quarter of the recruited participants (n=236) were previously part of the FAMILY Cohort, a prospective population-based study consisting of 46,001 participants in Hong Kong established to understand the determinants of physical, mental, and social well-being in Hong Kong^18^.

The HHGP-HK clinical study protocol was first approved by the Institutional Review Board of The University of Hong Kong/Hospital Authority Hong Kong West Cluster (HKU/HA HKW IRB) on 16 August 2021 with reference number UW21-549. HHGP-HK is registered with the HKU Clinical Trial Registry (HKUCTR) and is available for public access on www.HKUCTR.com with study identifier HKUCTR-2959. The study is sponsored by the Centre for Immunology & Infection (C2i) and was conducted as a single center study at the University of Hong Kong without any investigational product. The protocol is registered under ClinicalTrials.gov (study number NCT05174624). Three subsequent HKU/HA HKW IRB clinical research ethics review approval amendments were obtained to adapt to recruitment challenges due to the COVID-19 pandemic. Prior to launching the full study, a pilot study was conducted between 16 May 2022 and 13 June 2022 involving 106 participants (Table S1). The response and cooperation rates (Table S1) were comparable to those attained for random or population-based studies locally and internationally including pre-and peri-pandemic studies ^19^.

### 2.2 Study inclusion and exclusion criteria

Our study design was based on the definition of “health”, including both physical and mental well-being (Geneva: World Health Organization; 2020. Licence: CC BY-NC-SA 3.0 IGO), with the goal to maximize our ability to identify genetic and environmental associations with immune variability outside of a disease context. This was achieved by establishing a detailed list of inclusion and exclusion criteria ensuring the recruitment of volunteers with an immune system minimally perturbed based on a previously described definition of health from the Milieu Interieur study^2^ with some modifications made to adapt to the local population (e.g., Body Mass Index (BMI) category cut-offs for Asian population, local health authorities’ criteria for binge drinking), to the enhancement of study practices (e.g., inclusion for documented whitecoat hypertension), and to the evolving definition of “healthy” (e.g., use of statins and certain mental medications) as well as an increase in the age range to 79 years old. Briefly, donors could not have evidence of, or report a history of specific neurological or psychiatric disorders, or severe/chronic/recurrent pathological conditions. Other exclusion criteria included: recent (3 months) use of illicit drugs (including cannabis), recent (6 weeks) vaccine administration, and recent (6 months) use of immune modulatory agents. To avoid the influence of hormonal fluctuations in women during the peri-menopausal phase, only pre-or post-menopausal women were included. A full list of inclusion and exclusion criteria is listed in Table S2.

### 2.3 Healthy donor identification and recruitment

To identify and recruit the cohort we conducted a specific recruitment campaign which involved approximately 10,000 instant messages, 8,779 written invitations, and 5,307 phone calls between July 2022 and September 2023. Potential donors from the existing FAMILY Cohort^18^, as well as their affiliates (i.e. any individuals referred by a FAMILY Cohort participant), and the general Hong Kong public were targeted. Invitation pamphlets with the study contact information were sent to the residential addresses of potential participants of the FAMILY Cohort members, as well as distributed in public places and organizations. Interested individuals contacted our study researchers directly via mail, phone, email or instant messages, with potential participants approached by research assistants using a recruitment script to assess eligibility for the study.

This study consisted of two clinical visits: Enrolment (V0) and Inclusion (V1). All participants were first pre-screened through an interview conducted either via telephone or online to provide information such as year of birth, general health status, and vaccination history. Those screened as eligible for study participation were invited to the Enrolment Visit (V0). Individuals who refused to participate in the study were asked to provide basic descriptive information (e.g., sex and age) and reason for refusal (e.g., not interested, too busy) to assess participation rate and potential biases. Verbal consent was obtained prior to collecting personal information, including year of birth, vaccination history, and brief personal medical history.

From these initial contacts 2,146 questionnaire interviews and 2,230 physical examinations were conducted. A single clinical investigator applied the established inclusion and exclusion criteria during the recruitment period to ensure consistency in subject enrolment. As a result of the physical examinations, 628 clinical referrals were made for further potential health interventions.

Upon completing Enrolment visit (V0), participants were assessed to determine eligibility for the Inclusion visit (V1). Throughout the visits, participants completed a set of questionnaires (in-person or via online platforms), underwent clinical assessments, health apparatus investigations (electrocardiogram, dual-energy X-ray absorptiometry scan, and 7-day wearable fitness trackers), and were invited to provide biological specimen samples (e.g. blood, stool, and nasopharyngeal swabs). Enrolment visits (V0) were completed for 1,119 participants, from which 1,025 donors were included in the full study (V1) between 1 July 2022 to 14 September 2023. Table 1 highlights the study schedule and questionnaire assessment.

**Table 1.** Study schedule and questionnaire assessment.

| Assessment | Timepoint |  |
| --- | --- | --- |
|  | Visit 0 (Enrolment)<br>Day 0 | Visit 1 (Inclusion)<br>Median: 22 days |
| Informed consent | x |  |
| Questionnaire | x | x |
| Vital signs | x | x |
| Electrocardiogram (ECG) | x |  |
| Blood collection for Biochemistry | x |  |
| Blood collection for Haematology | x |  |
| Blood collection for Virology | x |  |
| Blood collection for TruCulture tubes |  | x |
| Blood collection for Immunophenotyping |  | x |
| Nasal swabs Specimens |  | x |
| Stool Specimens |  | x |
| Hair Specimens (n = 779) |  | x |
| Wearable devices (Fitbit) (n = 696) |  | x |
| Bone density scan (DXA scan) (n = 118) |  | x |
|  | n = 1,119 / 1,025 | n = 1,025 |

After consent was obtained, a unique study subject identifier was given to each participant. Pre-inclusion evaluation included the following assessments: (i) Vital signs and anthropometrics, (ii) Height, weight, BMI, (iii) Waist circumference, (iv) Tympanic temperature, (v) 2 Supine blood pressure readings (spaced 5 minutes apart), (vi) Resting heart rate, (vii) 12-lead electrocardiogram, (viii) Medical questionnaire to note personal medical history, substance use, vaccination history, concomitant medications, (ix) 25mL of blood collected for complete blood count (CBC), renal/liver function test (R/LFT), gamma-glutamyl transferase (GGT), thyroid stimulating hormone (TSH), uric acid, lipids, fasting glucose, C-Reactive Protein (CRP), infectious disease serology (i.e. HBV, HCV, HIV, HTLV I/II), and other viral serology (e.g. influenza/ COVID-19: for participants naïve to any COVID-19 vaccines), (x) Urine pregnancy test (for non-menopausal female participants), (xi) Full physical examination (conducted by Clinical Investigator). Resting heart rate was measured after resting for 5 minutes in a supine position and two readings were taken 5 minutes apart. If abnormal findings were observed that were deemed clinically significant, the Clinical Investigator determined the participant’s eligibility, and if required made referrals for further clinical management.

### 2.4 Biological sampling

Blood was collected by venepuncture performed by a registered nurse, or research personnel trained in phlebotomy under the supervision of a medical doctor/nurse. A Research Assistant was present throughout to double-check specimen labels and fill in the Specimen Collection documentation corresponding to the visit. The blood specimens for standard laboratory data analysis during the Enrolment visit (V0) or Inclusion visit (V1) were collected in specified Vacuette tubes and transported to an outsourced clinical testing laboratory within the same day of collection. During the Inclusion visit (V1) blood specimens (n=1025) for serology testing were collected in clotted blood tubes, for flow cytometry and PBMC isolation in Lithium Heparin Vaccuette® tubes, for genetic and protein analysis in EDTA tubes. All such V1 blood tubes were transported to the Centre for Immunology and Infection (C2i) at 18-25°C within the same day of collection before performing study analysis or storage. Additional blood samples were collected in Lithium Heparin Vaccuette® tubes and transferred to TruCulture® tubes within 30 minutes at the processing site of the research clinic. After incubation of the TruCulture® tubes at 37°C for 22 hours the samples were separated into supernatants and cell pellets and cryopreserved in dry ice at -80°C for storage.

Nasopharyngeal swabs (n=1025) were obtained bilaterally with sterile, dry swabs by the registered nurse or the Clinical Investigator in a designated isolated room equipped with IQAir Cleanroom Series filter. The designated research personnel was fully equipped with Personal Protective Equipment (PPE), including an isolation gown, non-sterile gloves, and a face shield. A Research Assistant was present throughout to double-check specimen labels and fill in the Specimen Collection documentation corresponding to the visit. Upon collection, all nasopharyngeal swabs were stored upright in a rack, and placed in a thermal bag for temporary storage before transport to the C2i for storage at -80°C.

Faecal samples (n=1025) were collected using the OMNIgene-GUT kit which is composed of a collection tube containing stabilising liquid, a spatula, and an instruction manual. Prior to the Inclusion visit (V1), participants were given the stool collection kit that was labelled with their respective study subject identifier to provide a faecal sample prior to their scheduled V1 visit. Participants were advised to put their own faecal specimens in sealed double bags before passing it to the research personnel at the clinical site, from where it was transported within the same day to the C2i for cryopreserving and storage at -80°C.

Approximately 100 strands or 50 mg of hair were cut from participants (n=548) as close as possible to their scalps, by a research nurse or a research assistant. The process started by positioning a piece of filter paper on an aluminium foil with a line indicating the end of hair closest to the scalp. A comb was used to partition the hair between the ears on the back of the head below the midline in the occipital area, which was selected for hair sample collection. Then it was cut with a pair of blunt tip scissors. If the hair was too short to be cut and clipped together, then the hair was cut directly into the foil. Hair samples were kept in a zip closable bag in a dry, dark environment at room temperature, away from direct sunlight until shipping to the School of Public Health, The University of Hong Kong for storage.

### 2.5 Wearable fitness trackers

A Fitbit Inspire 2 (Healthy Metrics Research, Inc. Fitbit, Inc., San Francisco, CA, USA) with instructions for use was provided to participants (n=903) to be worn on the wrist of the nondominant hand to collect 7-day health/ biometric and lifestyle data including daily step count, heart rate variability (HRV), and duration of sleep information. Participants were encouraged to wear the device continuously throughout the assigned 7-day period except for an hour each day during bathing or showering. Fitbit accounts were registered anonymously for managing health/ biometric and lifestyle data of participants. No identifiable personal information was used during the account registration process to ensure anonymity following the University of Hong Kong Code of Practice on personal data protection and Personal Data (Privacy) Ordinance (Cap. 486 of Laws of Hong Kong). Data was obtained from participants’ Fitbit accounts. Data from participants who had completed at least three consecutive nights of daily HRV measurements were considered for inclusion in the study. This criterion reduced the potential impact of day-to-day variations or abnormalities that could be present in a single day. After such selection, the data from 696 participants remained and mean values and standard deviations of each variable were calculated for further analysis.

### 2.6 DXA bones scans

Selected participants aged 50 or above (n=106) received dual-energy X-ray absorptiometry (DXA) scans (Explorer S/N 91075, Hologic Inc., Waltham, MA, USA) for lumbar spine and hip bone mineral density evaluation, and body composition determination. The DXA scan was conducted by the University of Hong Kong Centre for Sports and Exercise (CSE) at the B-Active fitness centre (https://www.cse.hku.hk/).

### 2.7 Case report forms

Detailed medical history and questions about socio-demographic, lifestyle and family health history were recorded in an electronic case report form via Clinical Management System and Qualtrics. It included information concerning family status, income, occupational status and educational level, smoking habits, alcohol intake, sleeping habits, depressive symptoms, family medical history and nutritional behavior and habits (for details, see supplementary material: case report forms). Participants provided the data detailed in Table 1 by completing the respective questionnaires administered by designated research personnel.

### 2.8 Milieu Interieur cohort data

To compare the HHGP-HK donors with an equivalent healthy donor cohort we used previously published data sets from the Milieu Interieur cohort, described elsewhere^2^. Briefly the study was approved by the *Comité de Protection des Personnes*-Ouest 6 on June 13^th^, 2012, sponsored by Institut Pasteur (Pasteur ID-RCB Number: 2012-A00238-35) and registered under ClinicalTrials.gov (study# NCT01699893). The data used in this study were formally established as the *Milieu Intérieur* biocollection (NCT03905993), with approvals by the *Comité de Protection des Personnes – Sud Méditerranée* and the *Commission Nationale de l’Informatique et des Libertés* (CNIL) on April 11, 2018. Donors gave written informed consent.

### 2.9 Statistical analysis

Statistical analysis was performed using the Open Source R Software^20^, version 4.3.0. The data curation and preprocessing were carried out using the tidyverse^21^ package (version 2.0.0), the broom.mixed^22^ package (version 0.2.9.5), and the rio package (version 0.5.29). Statistical graphics were generated using the ‘ggplot2’^23^ package, version 3.5.0 and ggpubr^24^ package, version 0.6.0. Principal component analysis (PCA) to identify potential outlier cases in our dataset was performed with the ‘FactoMineR’^25^ package version 2.9. Regression analyses were conducted using the lm function in R ‘stats’ package, version 4.3.0 and gamlss^26^ package, version 5.4-22. The GAMLSS package was employed using a normal distribution. The response variables are assumed to be normally distributed with age and sex and their interaction. The model is expressed as y∼age+sex+age:sex for the mean response and σ∼age+sex+age:sex for the sigma parameter, without any random effects. All the scatter plots generated using the ggplot2 package, were created using the ’LOESS’ method within the geom_smooth() function. Standardized regression coefficient estimates were obtained through z-score normalization and used to generate a forest plot using the ggforestplot package (version 0.1.0), along with their corresponding 95% confidence intervals. To account for multiple testing, the Benjamini-Hochberg method for false discovery rate (FDR) correction was applied to the p-values across all response variables for each of the predictor variables (age, sex, and their interaction) using the p.adjust() function. The geographical map was generated using the ‘sf’^27^ package, version 1.0-14. The comparison between the HHGP-HK donors and the Population Health Survey 2020-22 was performed using Chi-squared tests. Related heatmap visualization was generated using the ComplexHeatmap^28^ package (version 2.18.0).

## 3. Results

### 3.1 Establishment of a healthy donor cohort in Hong Kong

From July 1^st^ 2022 to September 14^th^ 2023, we recruited and sampled 1,025 well-defined healthy donors as defined by strict inclusion and exclusion criteria (Table S2) in Hong Kong (Figure 1a) across two sampling visits. Between the enrolment and inclusion visits, 151 donors were excluded and 22 withdrew from the originally pre-screened 1,198 donors (Figure 1b) due to different reasons (Table S3). The major exclusion criteria at this recruitment stage, which was after an initial screening interview, were BMI (n=28) outside of the defined range (18.5-27.0)^29^ or recent infection (n=26). In addition, certain autoimmune conditions were diagnosed during the medical visit and initial blood chemistry analysis including thyroid dysfunction (n=12), ulcerative colitis (n=1), and rheumatoid arthritis (n=1) leading to exclusion from the main cohort. Sampling of donors avoided temporal recruitment biases for age and sex (Figure 1c) and occurred at a clinic hosted at HKU B-Active centre of the University of Hong Kong located on the main island of Hong Kong, with donors coming from all territories of the wider Hong Kong region (Figure 1d). We compared socio-demographic information for our study subjects with the participants of the Population Health Survey 2020-22 conducted by the Department of Health, the Government of the Hong Kong Special Administrative Region (https://www.chp.gov.hk/en/features/37474.html). From this analysis we observed that the HHGP-HK donors had generally healthier behaviours (e.g. increased exercise and activity, less smoking) (Figure S1a) and biological laboratory measures (Figure S1b) as expected from our strategy to specifically select healthy individuals.

**Figure 1.**
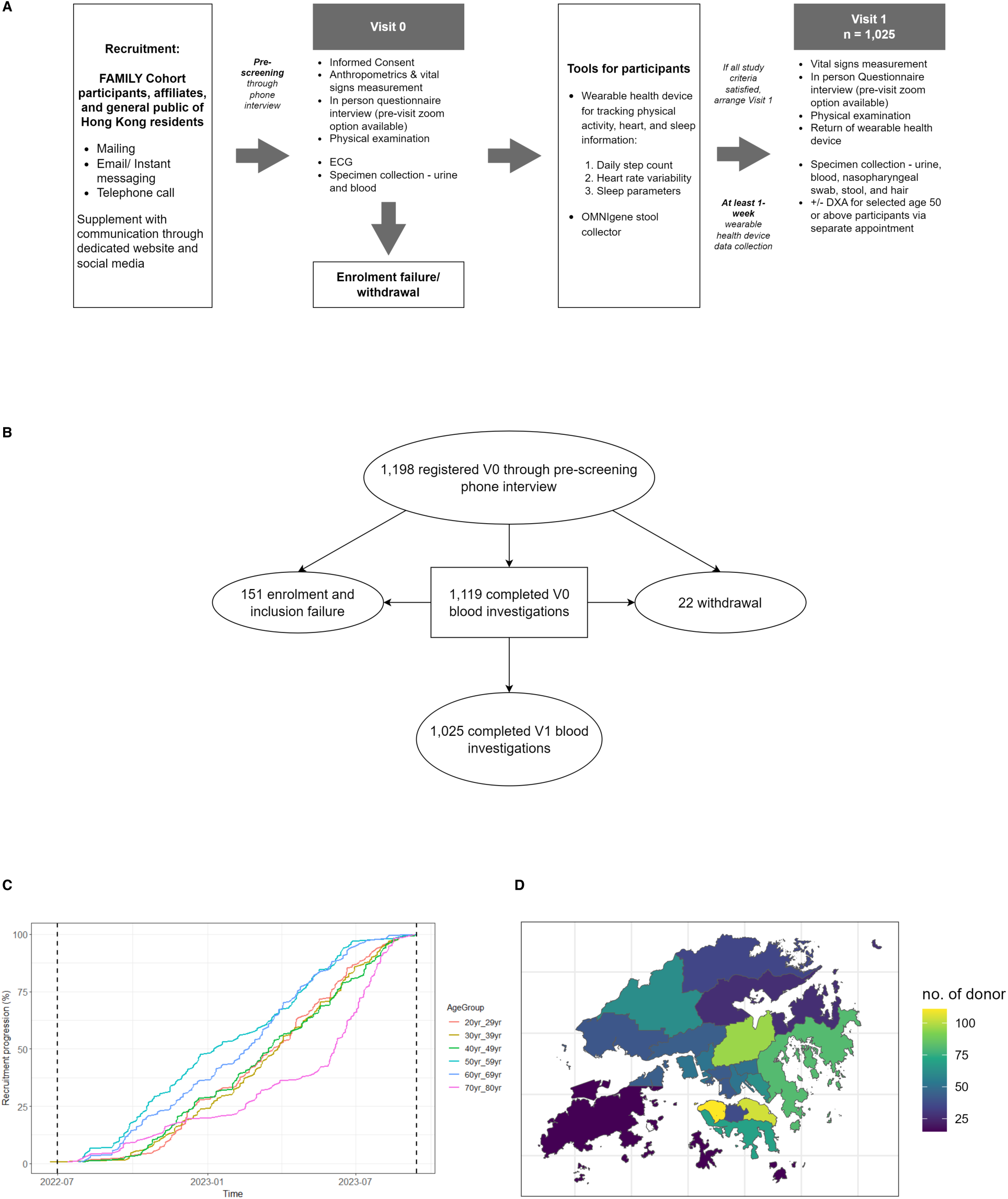
Schematic representation of donor recruitment for the HHGP-HK study. (A) Outline of donor recruitment strategy, Visit 0 assessment, wearable inclusion and Visit 1 biological sampling. (B) Breakdown of donor recruitment from V0 to V1. (C) Timeline of donor recruitment per age decade (D) Geographical distribution across the Hong Kong region of recruited donors.

This major recruitment effort succeeded in attaining our main objective, which was the establishment of a healthy donor cohort to allow the study of genetic and environmental associations with immune variability outside of a disease context. To address these questions, we collected extensive demographic and medical history information by questionnaire, as well as multiple biological samples described in Table 1.

### 3.2 Sex and age effects on clinical laboratory measures in HHGP-HK donors

To establish healthy reference range values for the HHGP-HK cohort we measured multiple biochemical, hematologic and serologic phenotypes in the blood using standardized laboratory assays and clinical tests. To assess the quality of the data sets we examined for well-known age and sex effects, as well as assessing age:sex interactions, through linear regression models. Out of 66 variables tested we identified a significant effect (FDR p < 0.05) of age on 37 variables, of sex on 48 variables, and their interactions on 36 variables (Figure 2a). As an example, we illustrate serum creatinine, an important metabolite that showed the strongest sex differences, with significantly higher levels in males compared to females (Figure 2b,c) as previously described^30^. Blood pressure, alkaline phosphatase (ALP) and sodium levels showed the highest increases with age, while IgM and albumin (ALB) showed the strongest declines (Figure 2a). For sex differences, proteins such as hemoglobin, hematocrit, urea, and urate, were significantly higher in males and showed significant sex:age interactions (Figure 2a). In contrast HDL, phosphorous, IgM and platelets were the variables that were most significantly higher in females, of which phosphorous and IgM showed significant sex:age interactions (Figure 2a).

**Figure 2.**
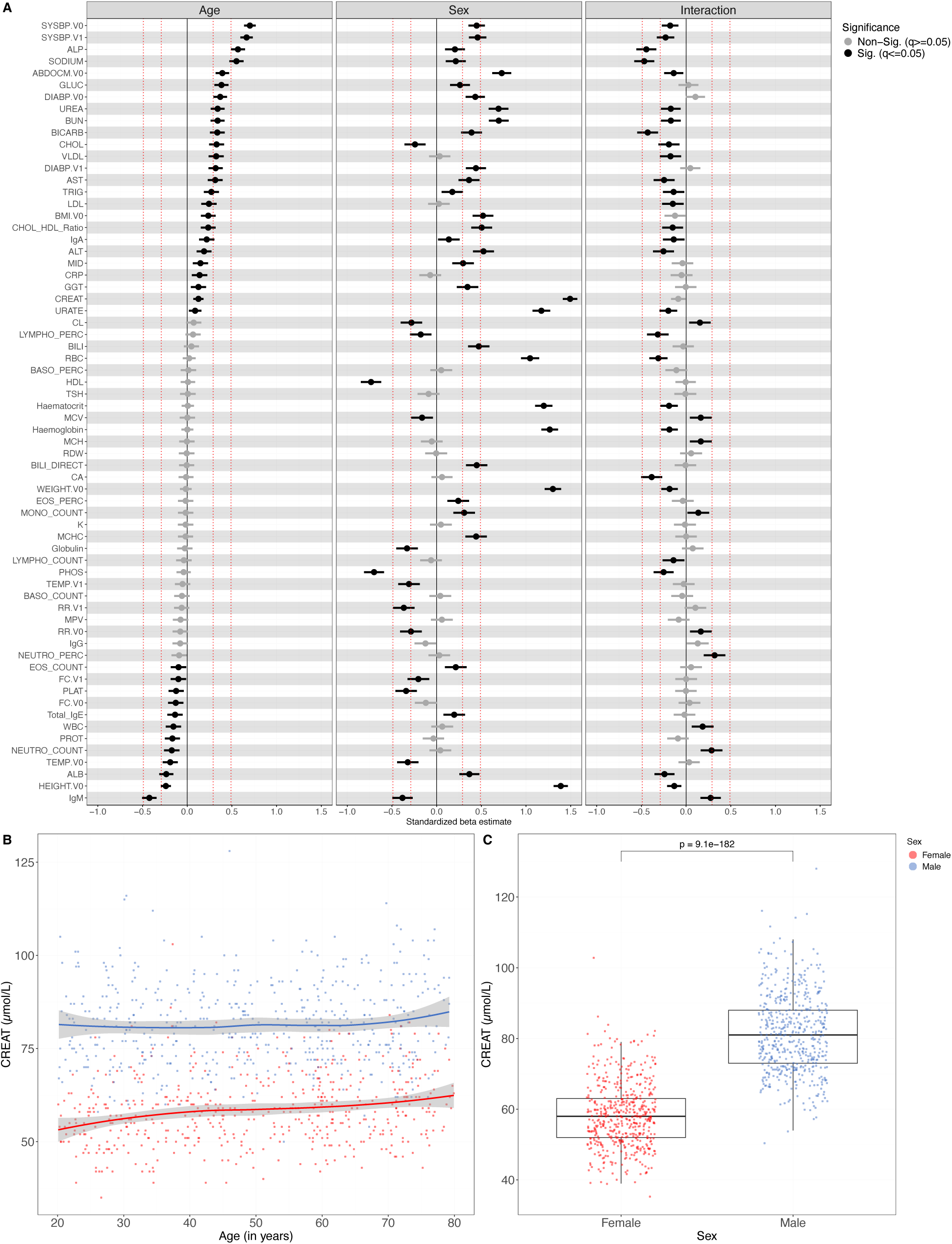
Age, sex, and age:sex interaction effects on clinical laboratory measures. (A) Forest plots of effect sizes (Standardized regression coefficient estimate) of age, sex, and age:sex interactions on the mean of 66 clinical laboratory measures in the HHGP cohort. (B) Blood plasma levels of creatinine (umol/L) as a function of age and (c) sex. Error bars represent 95% confidence intervals. T test, Spearman regression, (n=1025).

### 3.3 Comparison of age and sex effects on clinical laboratory measures between healthy Asian (HHGP-HK) and European (Milieu Interieur) donors

Directly comparing human cohorts analyzed in different locations and at different times can be challenging due to batch effects inherent in many biological measures^31^. To overcome this challenge, while making a first comparison with another healthy cohort of different ancestry we first analyzed age and sex effects on clinical laboratory measures between HHGP-HK and MI donors^2^. To ensure robust comparisons we compared donors from the same age range (20-69 years) and downsampled at random the MI cohort to match the size of HHGP-HK within this age range (n=856), while keeping a balanced sex ratio. Overall, the age effects were remarkably consistent between the two cohorts with blood pressure, sodium, ALP, and glucose among the many variables significantly increasing with age in both cohorts, while albumin, total protein concentrations, and neutrophil counts were among the variables significantly decreasing with age (Figure 3a). Some analytes showed discordant patterns between the two cohorts, notably CRP which had higher levels in MI and showed a significant age:sex interaction (Figure 3a). Most of the analytes showed consistent sex differences but with stronger effect sizes in either cohort (Figure 3d), for example gamma-glutamyltransferase (GGT) had a stronger male bias in MI and abdominal circumference a stronger male bias in HHGP-HK (Figure 3a). As an example we illustrate blood albumin levels which were consistently higher in males of and with a moderate age decline in both cohorts (Figure 3b, c).

**Figure 3.**
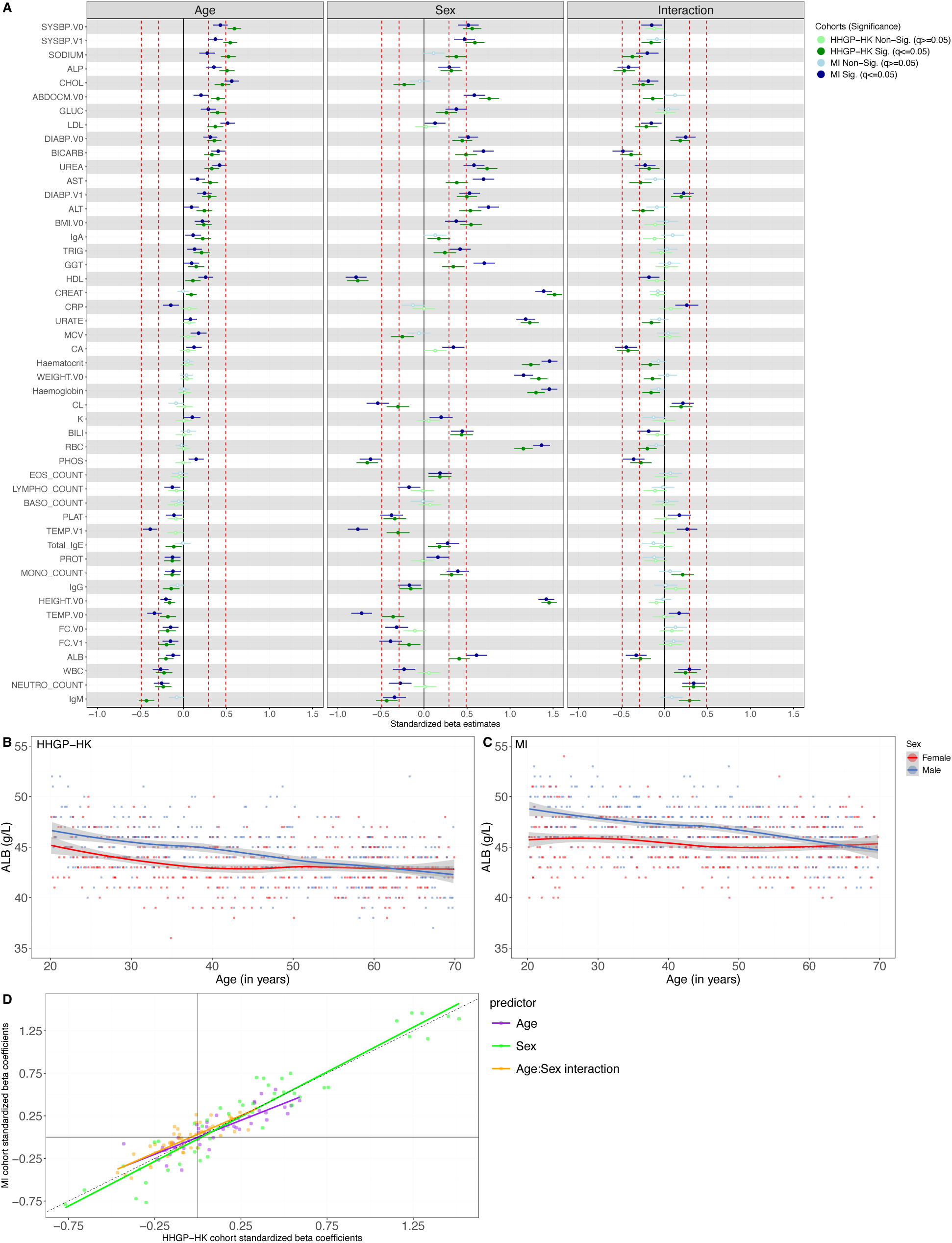
Age, sex, and age:sex interaction effects for the age and sex matched HHGP-HK and MI cohorts. (A) Effect sizes (Standardized regression coefficient estimate) of the effects of age, sex, and age:sex interactions on 49 clinical laboratory measures in the HHGP and MI matched cohorts. Blood levels of albumin (g/L) in female and male donors of (B) HHGP and (C) MI donors. (D) Correlation plot of standard beta coefficients for age, sex, and age/sex interactions between HHGP and MI cohorts. (n=856).

Such differences between males and females can be a result of biological effects (sex), as well as potential gender effects, meaning the different socio-cultural impacts from living in a specific society as a man or a woman. Disentangling sex from gender effects on biological phenotypes can be challenging, but cross comparisons between diverse healthy cohorts where gender may have different local cultural impacts can be a first step to address this question. In line with this concept we looked for variables with a significant age:sex interaction in both cohorts, that also had consistent sex differences in mid-life suggesting changes due to menopause. We identified soluble blood factors and immune cell populations that showed the same opposing pattern between female and male donors throughout life, with the change occurring around the age of 50 (Figure 4). Specifically, blood cholesterol (Figure 4a, b) calcium (Figure S2a, b), and LDL (Figure S2c, d) were higher in men early in life, and higher in women later in life. Showing the opposite pattern blood neutrophil counts (Figure 4c, d) and white blood counts (WBC) (Figure S2e, f) were higher in women early in life, but higher in men later in life. As examples of phenotypes with age consistent sex differences platelets were consistently significantly higher in female donors (Figure 4e, f), and red blood cells (RBCs) were higher in male donors across all ages in both cohorts (Figure S2g, h). This raises the hypothesis of sex hormonal effects on neutrophil and white blood cells counts, but not platelets, a hypothesis we will test in future studies.

**Figure 4.**
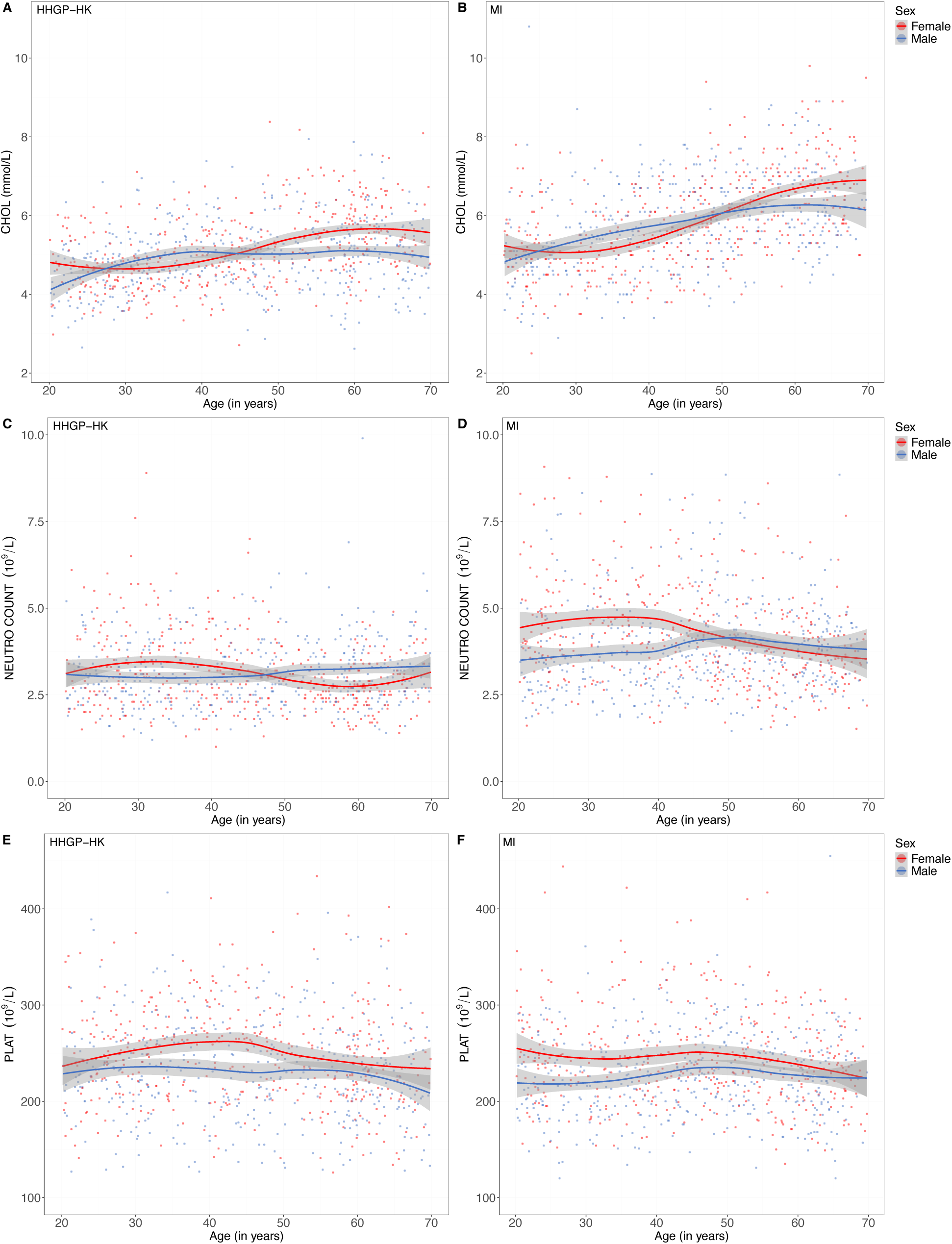
Significant age:sex interactions on blood phenotypes in HHGP-HK and MI. Blood concentrations for females and males of HHGP-HK for (A) total cholesterol (mmol/L), (C) neutrophil counts (10^9^/L), (E) platelet counts (10^9^/L) and for MI donors (B) total cholesterol (mmol/L), (D) neutrophil counts (10^9^/L), (F) platelet counts (10^9^/L). (n=856).

We also wanted to test whether clinical laboratory measures were significantly more variable either with increased age, or between female and male donors in both cohorts. To test this, we applied Generalized Additive Models for Location, Scale, and Shape (GAMLSS) to our data sets. In contrast with more conventional linear models, GAMLSS allows for accomadating skewness and heteroscedasticity within the data, consequently allowing to model and statistically compare variances (and other distributional moments) rather than just means^26^. We modelled the variance for all clinical laboratory measures for each cohort as a function of age, sex, and age:sex interactions (Figure S3). For the effects of age on variance most analytes were consistent between the two cohorts with a notable exception of total IgE levels, the variance of which decreased with age in HHGP-HK but increased in MI (Figure 5a, b & Figure S3). Variance of IgE levels was also significantly higher in male donors of both cohorts, but the differential age effects in MI resulted in a significant age:sex interaction (Figure S3). These observations may reflect different environmental allergic exposures between the two locations. Regarding sex effects on variance, as for age most variables were consistent between the two cohorts with the exceptions of three liver enzymes; alanine transaminase (ALT) (Figure 5c, d), gamma-glutamyl transferase (GGT) (Figure 5e, f), and aspartate aminotransferase (AST) (Figure 5g, h). All 3 enzymes showed higher variance between male and female donors in MI as compared to HHGP-HK (Figure 5 and Figure S3). These enzymes have previously been reported to be lower in Asian populations^32^, and the higher levels as well as higher variance in Europeans may reflect either genetic or environmental (eg diet, alcohol consumption) effects which will be tested in future studies.

**Figure 5.**
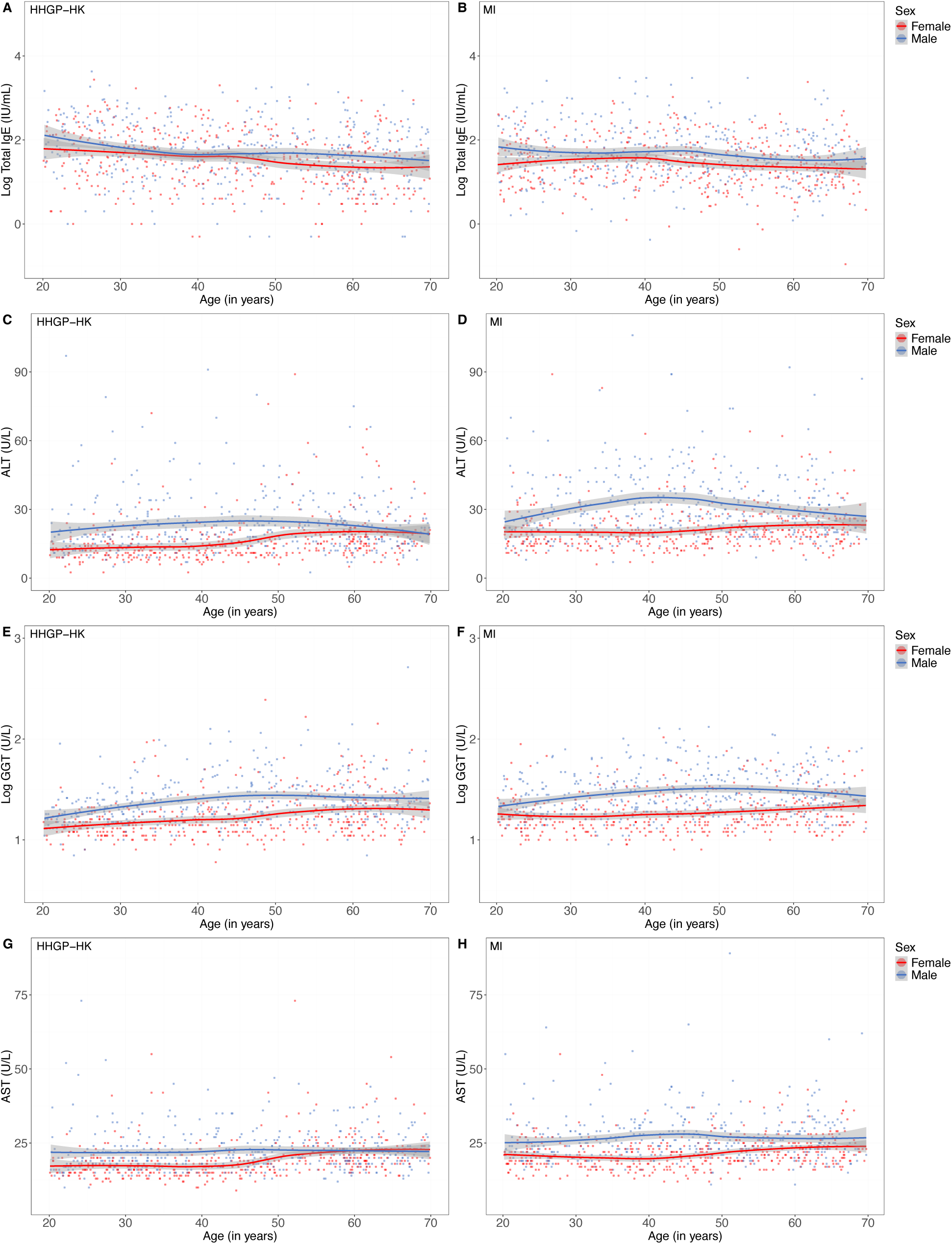
Blood phenotypes with different variance between HHGP-HK and MI. Blood concentrations for females and males of HHGP-HK for (A) log total IgE (IU/mL), (C) ALT (U/L), (E) log GGT (U/L) (G) AST (U/L) and for MI donors (B) log total IgE (IU/mL), (D) ALT (U/L), (F) GGT (U/L), (H) AST (U/L). (n=856).

### 3.4 Comparison of biomarker health scores between healthy Asian (HHGP-HK) and European (MI) donors

While individual biological measures are useful for specific diagnosis, combined biomarker scores are growing in popularity for health monitoring. An advantage of combined biomarker scores over individual measures is that they can capture physiological processes more broadly, and with the use of normalized scores are more resistant to batch effects and differences in technical measures that can occur between labs. With this in mind we calculated different biological scores on the HHGP-HK cohort. We first derived the metabolic score; a quantitative measure used to assess an individual’s metabolic health by combining abdominal circumference, systolic blood pressure, diastolic blood pressure, triglyceride, HDL, and fasting glucose^33^. We used cut-off measures previously adapted to a Hong Kong population to calculate index values, from which 14.73% had a score of 2 (n = 151), 6.24% a score of 3 (n = 64), 1.85% a score of 4 (n = 19), and 0.39% a score of 5 (n = 4) (Figure 6a, b). An increased score was strongly associated with age (p = < 0.0001), but not with sex (p = 0.07), although the age:sex interaction was significant (p = 0.02). The metabolic score was also higher in active smokers as previously described^2^.

**Figure 6.**
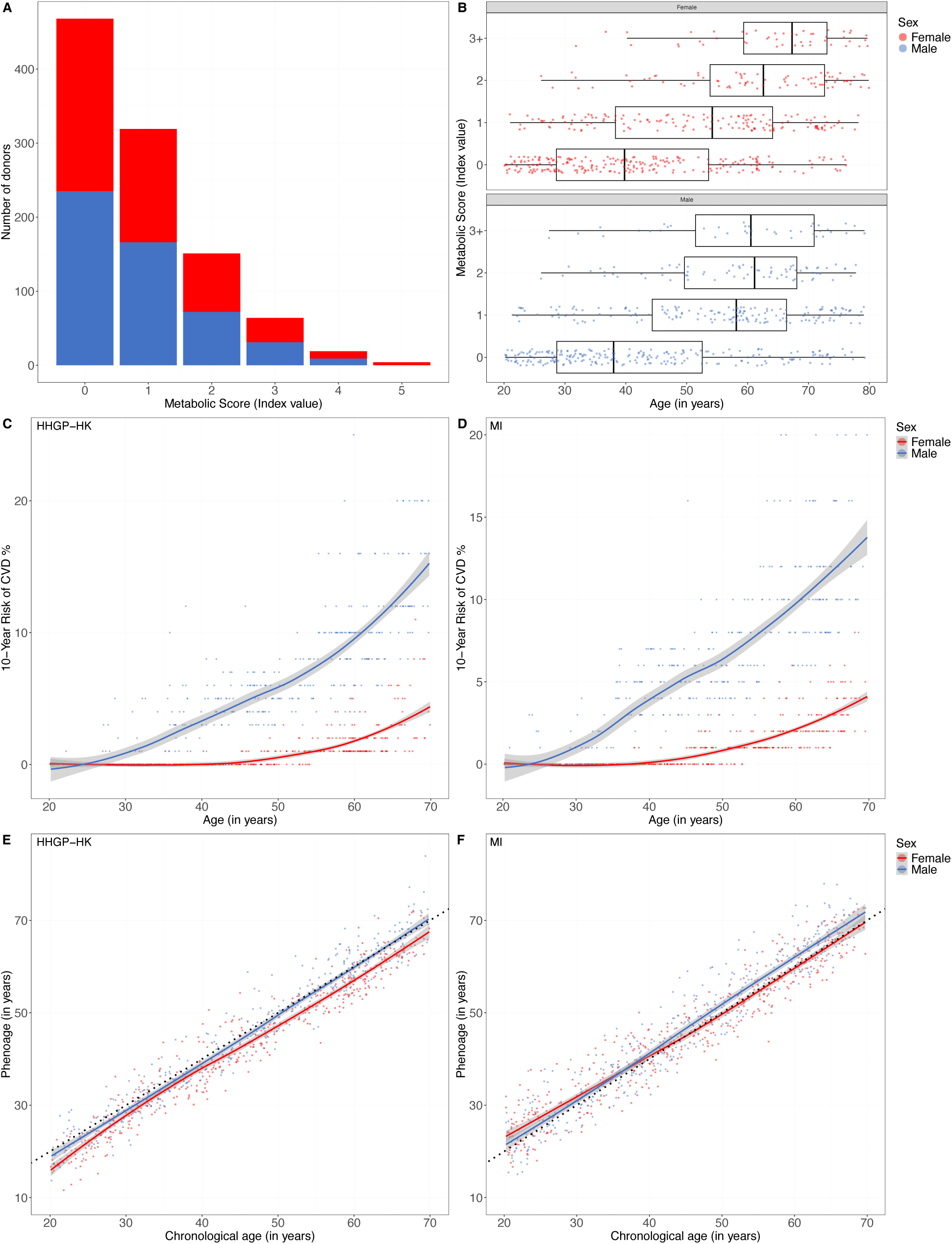
Health scores comparisons between HHGP-HK and MI. (A) Metabolic score categories for HHGP donors as function of sex and (B) age (n=1025). Framingham 10-Year Risk Score for (C) HHGP-HK and (D) MI. Phenoage and chronological age correlations for (E) HHGP-HK and (F) MI. (n=856).

We also calculated the Framingham 10-Year Risk Score for both HHGP-HK and MI^34^. This is a tool used to estimate an individual’s risk of developing cardiovascular disease (CVD) within the next ten years based on the following factors: age, total cholesterol, HDL, systolic blood pressure, treatment for hypertension, and smoking status. We observed highly consistent patterns between both cohorts with a significantly higher risk for men from both countries, and a shaper increase with age for males (Figure 6c, d).

Lastly, we calculated the PhenoAge score, a measure of biological age based on blood levels of albumin, creatinine, fasting glucose, CRP, mean corpuscular volume (MCV), ALP, % lymphocytes and white blood counts (WBCs)^35^. This score was originally trained on a DNA methylation data set and reflects biological age. In HHGP-HK donors the PhenoAge score correlated extremely closely with chronological age (R^2^ = 0.96, p < 0.001) for both males and females (Figure 6d). However, when comparing to the expected age regression line HHGP-HK females were consistently below the line showing lower biological age than chronological age. This raises two interesting hypotheses, either the PhenoAge score which was developed on Western populations is not adapted to Asians, or that females recruited in the HHGP-HK study have lower signs of biological aging. With the generation of future data sets we will test these intriguing hypotheses while integrating with immune phenotypes and functional assays.

### 3.5 Covid-19 infection and vaccination status in HHGP-HK donors

A major challenge for implementing the HHGP-HK study was the ongoing Covid-19 pandemic, with the SARS-CoV-2 Omicron wave occurring in Hong Kong in December 2021 resulting in acute infectious cases remaining prevalent during the pilot study. To study the impact of the pandemic on our cohort we included multiple questions related to Covid-19 in the CRF questionnaire. From this we identified that 56.3% of the cohort reported a confirmed infection (either PCR or antigen test) which is within the estimated rates in the overall Hong Kong population^36^. Main symptoms reported included classical ones such as fever (31.4%), coughing (35%), and sore throat (35.5%). Of the reported infections 56% were classified as moderate, 0.3% severe, and no critical cases. 34.1% of donors reported post-infection symptoms indicative of Long Covid^37^, which is also within the range reported in the overall population^36^. For vaccination, the majority of the cohort (97.2%) received the full regimen (3 doses), with the main vaccine types including CoronaVac (34.6%), Pfizer mRNA (65.1%), and Moderna (0.3%). Assessing vaccine type uptake showed a strong age (but not sex) association with younger donors significantly more likely to have received an mRNA vaccine (Figure S4a), while older donors more likely to have received CoronaVac (Figure S4b). This may reflect the earlier availability and rollout of the CoronaVac vaccine to elderly donors in Hong Kong. Future analysis will integrate this data with SARS-CoV-2 specific serology and T cell responses to try and better understand factors contributing to variability in the immune responses differentially induced by these two vaccine types.

Given the widespread challenges caused by the Covid-19 pandemic we also assessed the mental health of our donors through a series of questions. From these responses we calculated a generalized anxiety disorder (GAD-7) score^38^ and the PHQ-9 Patient Health Questionnaire-9 (PHQ-9) score which are tools for screening, diagnosing, monitoring, and measuring the severity of depression. While overall both the GAD-7 (Figure S5a) and PHQ-9 (Figure S5b) scores were relatively low, some donors (6%) reported anxiety or depression levels above healthy levels. Assessment of both scores with age showed a significant association with higher reports of both anxiety and depression scores in younger donors for both male and female donors (Figure S5). Future analysis will assess how such mental health issues may be associated with immune responses.

### 3.6 Activity measures from a wearable fitness device in HHGP-HK donors

Given the importance of physical activity for general health, and recent developments of noninvasive tracking devices, we offered our donors the possibility to wear a FitBit Inspire 2 after study inclusion, during the week before or after the biological sampling visit. For FitBit monitoring 903 donors participated, from which after quality controls we examined data from 696 donors among which there was no age or sex biases. From these measures we examined total daily steps and observed a significant sex difference (p < 0.0001), with males recording greater number of steps, as well as a significant non-linear age effect with the number of steps peaking around 60 years old (Figure 7a). Notably average daily steps were above the general recommendation of 10,000 steps (https://www.10000stepsaday.hk/?lang=en) for all age groups studied. Minutes of deep sleep (calculated based on movement, HRV, and sleep staging using FitBit algorithms), another important indicator of good health, was significantly different between males and females (p=0.002) and declined significantly with age for both sexes (Figure 7c). Utilizing FitBit algorithms we could also extract information on calories, overall distance, minutes asleep and awake, and future work will integrate these factors with different immune phenotypes. We also determined heart rate variability (HRV) by measurement of the root mean square of successive differences (RMSSD) between heartbeats

**Figure 7.**
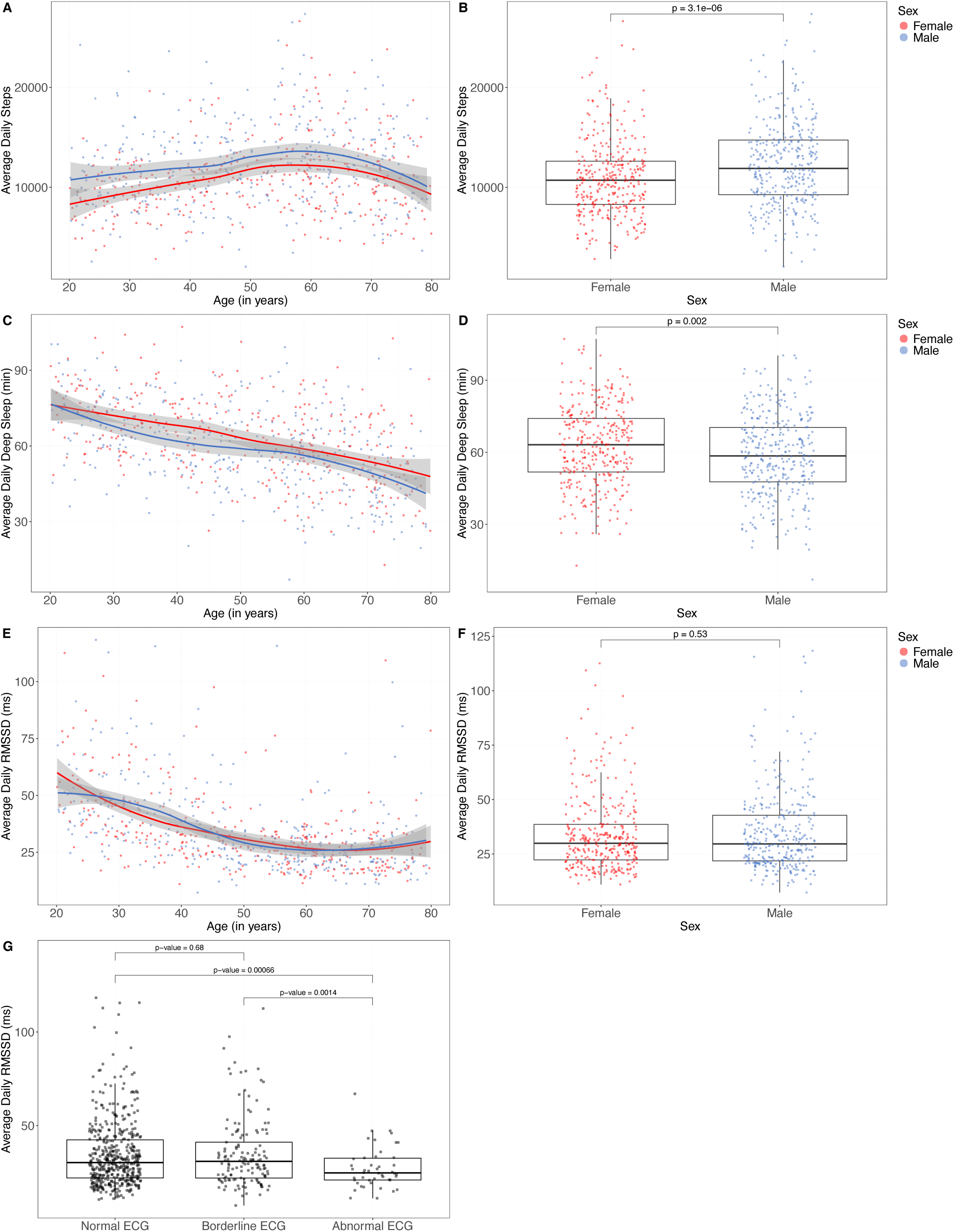
Physical activity from health tracking monitor in HHGP-HK donors. Average daily steps as measured by a wearable device in HHGP-HK as a function of (A) age and (B) sex. Average daily minutes of deep sleep as measured by a wearable device in HHGP-HK as a function of (C) age and (D) sex. Average daily RMSSD as measured by a wearable device in HHGP-HK as a function of (E) age and (F) sex (n=696). (G) Comparison of heart rate variability (HRV) between HHGP-HK donors with normal ECG, borderline ECG, and abnormal ECG. (n=672).

(Figure 7e). The majority of donors were within commonly reported reference ranges (20-70 milliseconds)^40^. No sex differences were observed for HRV with RMSSD though it did decline significantly with age as expected^40^. To test with more conventional heart electrical activity measurements, we stratified donors based on their Electrocardiogram (ECG) test in three groups; normal, borderline, and abnormal (Figure 7g). The ECG abnormal group had a significantly lower RMSSD value compared to the other two groups, demonstrating the capabilities of such wearable devices for health monitoring. Future studies will integrate these measures with diverse immune phenotypes to assess how such factors may influence immune variability.

## Discussion

We describe here the establishment of a unique cohort of healthy donors of East Asian ethnicity living in Hong Kong. We focused on well-defined healthy donors to further understand genetic and environmental determinants of biological variability outside of disease influences. This approach was pioneered in the MI cohort to identify novel genetic and environmental determinants of circulating immune cells^13^ and responses to microbial stimulation^41,42^. However MI was restricted to donors of Western European ancestry living in a single region in the West of France^2^ which is inherently restricted in terms of human and geographical diversity. The establishment of the HHGP-HK cohort is an effort to address these limitations and provide a better understanding of biological variability in an Asian population.

As a first test of our initial data sets from clinical laboratory measures we focused on age and sex effects, as our cohort is well powered to assess these factors which have widespread impacts on health. We also made a first comparison with healthy donors from the MI cohort which showed overall consistently similar age and sex effects on clinical blood measures. There were some interesting exceptions, for example CRP a commonly used inflammation marker, was higher in the blood of healthy French donors compared to Hong Kong. This is consistent with reports that CRP is generally lower in individuals of Asian ancestry^43^. Blood urea also showed interesting sex differences, with levels in MI females increasing with age in contrast to HHGP-HK females which remained significantly lower than males at all ages. These differences may reflect different diets between the two cohorts which will be investigated in future studies. More subtle differences were observed between the two cohorts when examining variance of the markers, as opposed to mean levels. This revealed consistently more variable liver enzymes (ALT, AST, GGT) in MI as compared to HHGP-HK donors. These markers are routinely used as biomarkers of liver physiology and the differences observed may reflect diet or alcohol consumption, or even underlying ethnic differences as suggested by previous studies^44^.

Indeed there is an increasing trend to move from the use of single biomarkers, which may be more subject to either inter-lab or intra-individual variability, to multi-analyte biomarker scores^45^. We applied two such well-known scores, the Framingham 10-Year Risk Score^34^ and PhenoAge^35^ to HHGP-HK and MI. While the Framingham score showed remarkably consistent results in both cohorts, PhenoAge aged the Asian and European donors differently. These results suggest that such aging and health prediction scores require both modification and validation in populations of different ancestries and locations, an objective that will be a future goal of this project.

One strength of comparing biological analytes in well controlled healthy cohorts stratified by age and sex from diverse regions is the ability to confirm significant age:sex interaction effects. Many normal physiological processes begin to decline in mid-life , a period of significant hormonal changes as reflected by the menopause, but we still lack a complete understanding of these aging mechanisms. In examining age:sex effects in both HHGP-HK and MI donors we observed significant mid-life changes in female donors for both circulating immune cells (neutrophils and white blood cells) and plasma lipids (LDL, cholesterol). The occurrence of these changes in midlife specifically in females suggests a strong implication of sex hormones, a hypothesis we will test, which if confirmed may have implications for the use of hormone replacement therapy ^47^.

Novel aspects of the HHGP-HK cohort include the use of wearable devices to track exercise and sleep, and assessment of mental health, both of which revealed significant age and sex differences. Future studies will focus on integrating these important aspects of health with immune phenotypes to better understand how they may interact. Some limitations of our study include a single recruitment site and a single sampling timepoint. Indeed longitudinal sampling is increasingly appreciated for understanding aspects of intra-individual biological variability, and the availability of complementary cohorts may allow for cross validation of specific findings . In summary the establishment of the HHGP-HK cohort has established foundations for better understanding immune variability in an Asian population to guide the development of future precision immunological strategies.

## Data Availability

All data produced in the present study are available upon reasonable request to the authors

## Acknowledgments

This study was funded by a grant from InnoHK, an initiative of the Innovation and Technology Commission, the Government of the Hong Kong Special Administrative Region. We also acknowledge and thank the FAMILY Cohort including Cynthia Yau, Annis Wong, Wong Hoi Wa for clinical study support and providing access for recruitment. The work is being submitted to the University of Hong Kong for the award of the degree of Doctor of Medicine. The cohort design was originally inspired by the Milieu Interieur Consortium, a project funded by French government’s Invest in the Future programme (reference ANR-10-LABX-69-01). We thank staff from HKUMed and the Centre for Sports and Exercise for facilitating the implementation of the project. Lastly we genuinely thank all of our donors for their active participation in this study.

The Milieu Intérieur Consortium¶ is composed of the following team leaders: Laurent Abel (Hôpital Necker), Andres Alcover, Hugues Aschard, Philippe Bousso, Nollaig Bourke (Trinity College Dublin), Petter Brodin (Karolinska Institutet), Pierre Bruhns, Nadine Cerf-Bensussan (INSERM UMR 1163 – Institut Imagine), Ana Cumano, Christophe D’Enfert, Ludovic Deriano, Marie-Agnès Dillies, James Di Santo, Gérard Eberl, Jost Enninga, Jacques Fellay (EPFL, Lausanne), Ivo Gomperts-Boneca, Milena Hasan, Gunilla Karlsson Hedestam (Karolinska Institutet), Serge Hercberg (Université Paris 13), Molly A Ingersoll (Institut Cochin and Institut Pasteur), Olivier Lantz (Institut Curie), Rose Anne Kenny (Trinity College Dublin), Mickaël Ménager (INSERM UMR 1163 – Institut Imagine), Frédérique Michel, Hugo Mouquet, Cliona O’Farrelly (Trinity College Dublin), Etienne Patin, Antonio Rausell (INSERM UMR 1163 – Institut Imagine), Frédéric Rieux-Laucat (INSERM UMR 1163 – Institut Imagine), Lars Rogge, Magnus Fontes (Institut Roche), Anavaj Sakuntabhai, Olivier Schwartz, Benno Schwikowski, Spencer Shorte, Frédéric Tangy, Antoine Toubert (Hôpital Saint-Louis), Mathilde Touvier (Université Paris 13), Marie-Noëlle Ungeheuer, Christophe Zimmer, Matthew L. Albert (Octant Biosciences), Darragh Duffy§, Lluis Quintana-Murci§,

¶ unless otherwise indicated, partners are located at Institut Pasteur, Paris

§ co-coordinators of the Milieu Intérieur Consortium

Additional information can be found at: http://www.milieuinterieur.fr

## Supplementary Materials

**Figure S1.**
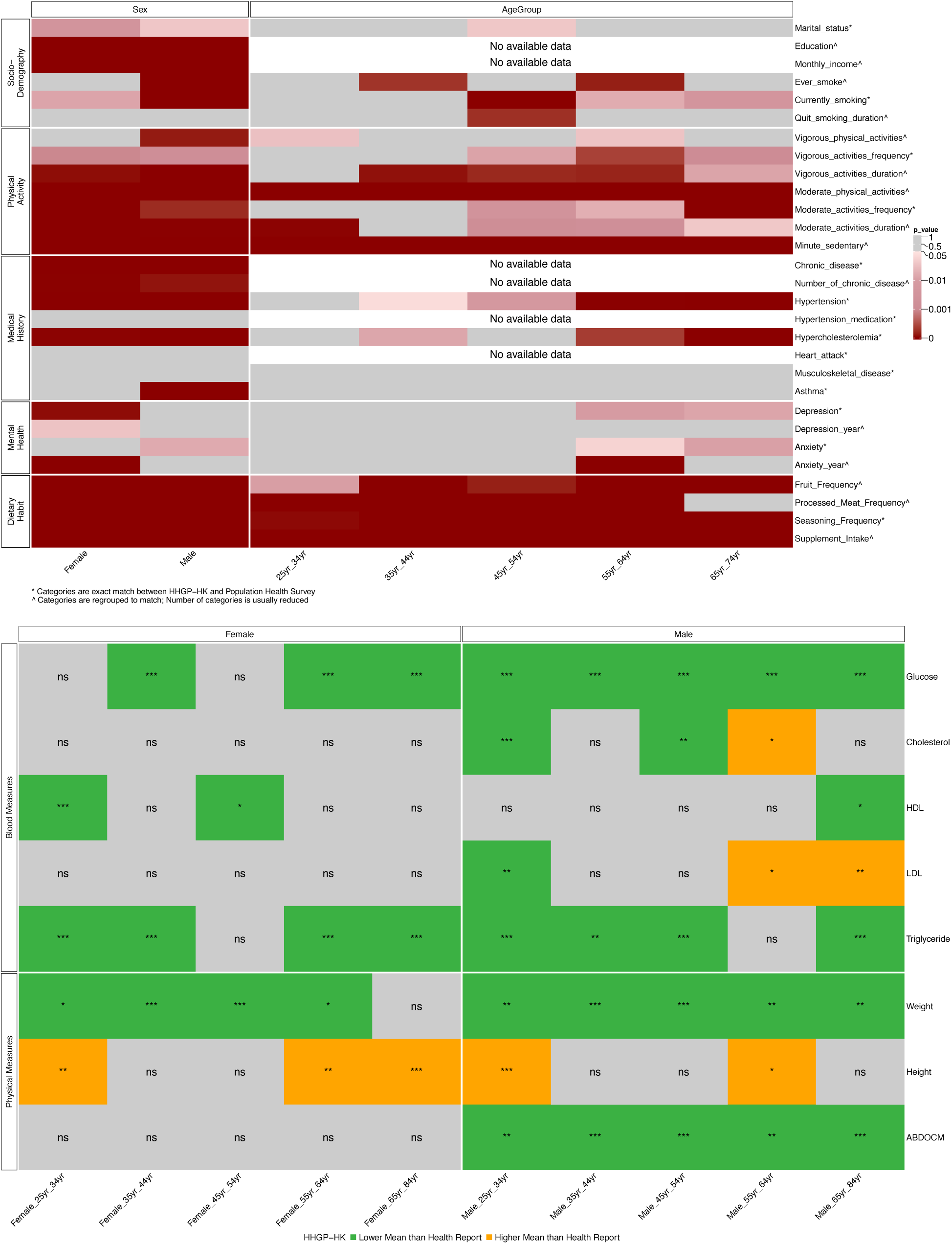
Comparison of HHGP-HK donors with Population Health Survey 2020-22. (A) Heat map of lifestyle and behaviour (B) Heat map of shared clinical laboratory measures. (n=1025)

**Figure S2.**
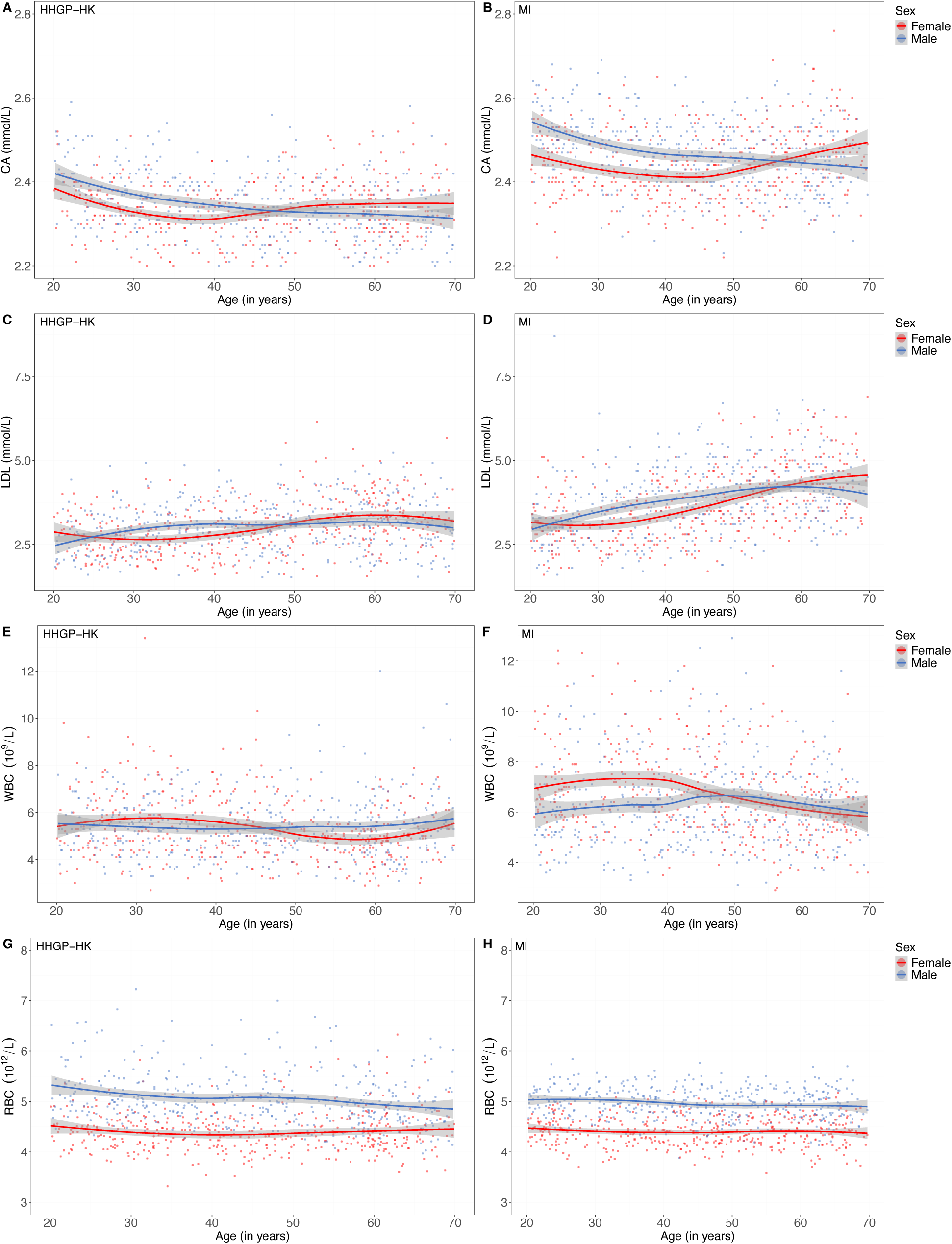
Significant age:sex interactions on blood phenotypes in HHGP-HK and MI. Calcium for (A) HHGP-HK and (B) MI, LDL for (C) HHGP-HK and (D) MI, White blood counts for (E) HHGP-HK and (F) MI, Red blood counts for (G) HHGP-HK and (H) MI (n=856).

**Figure S3.**
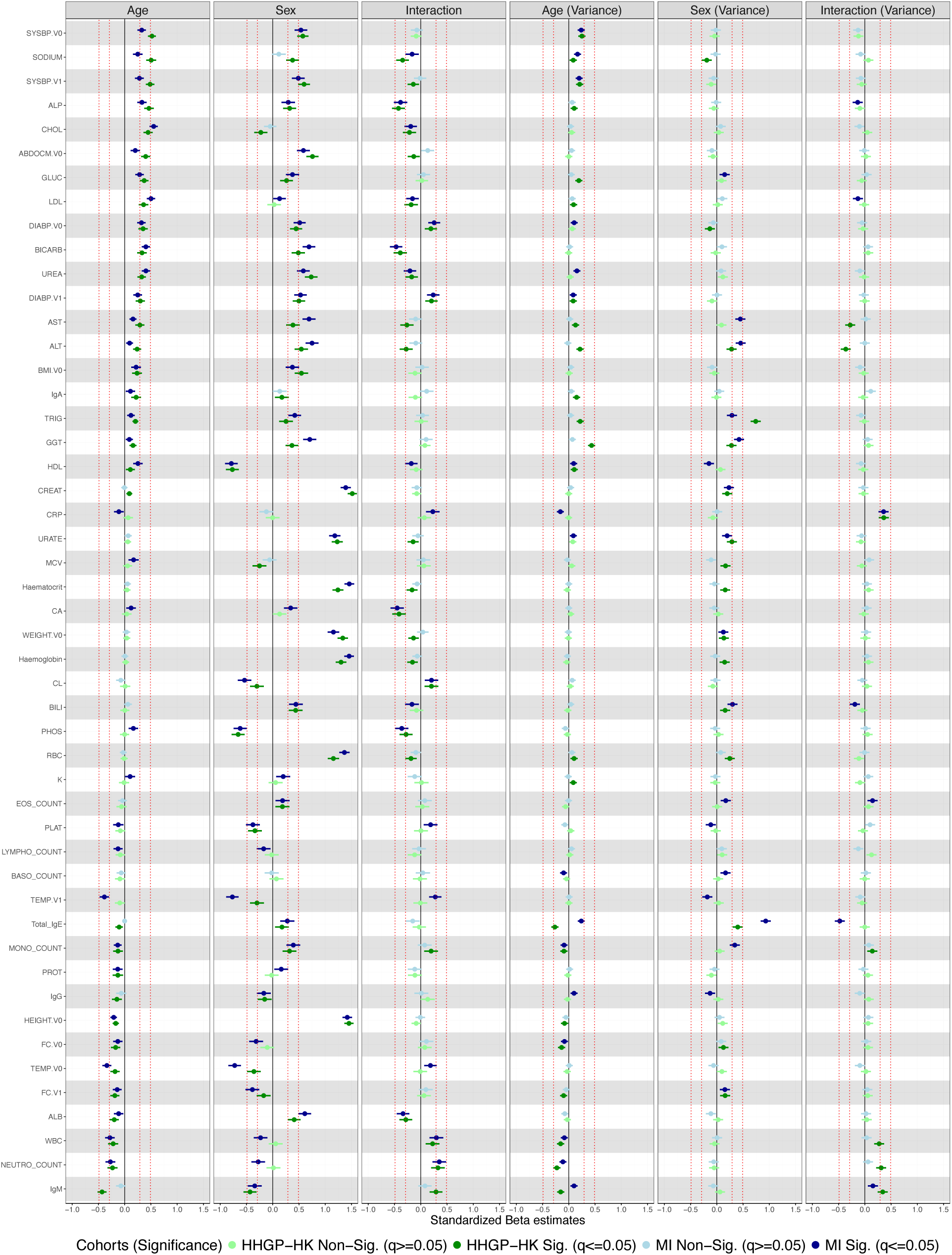
Age, sex, and age:sex interaction effects on variance of clinical laboratory measures for the age and sex matched HHGP-HK and MI cohorts. Forest plots of the effect sizes (Standardized regression coefficient estimate) of age, sex, and age:sex interactions on the variance of 49 clinical laboratory measures in the HHGP and MI cohorts. Error bars represent 95% confidence intervals. (n=856).

**Figure S4.**
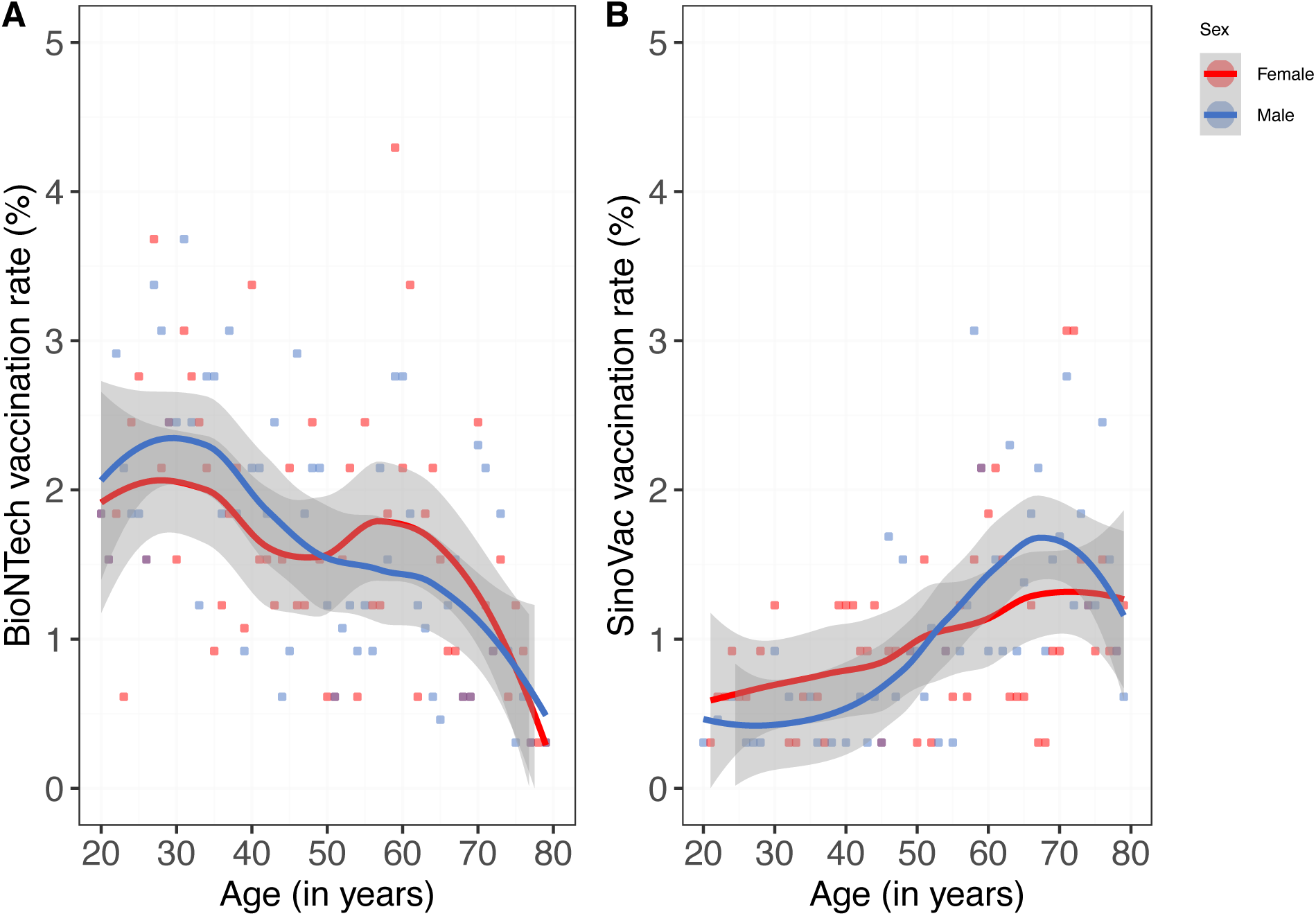
Age and sex distribution of vaccines in HHGP-HK donors. (A) mRNA and (B) CoronaVac vaccination rates as a function of age and sex in HHGP-HK donors.

**Figure S5.**
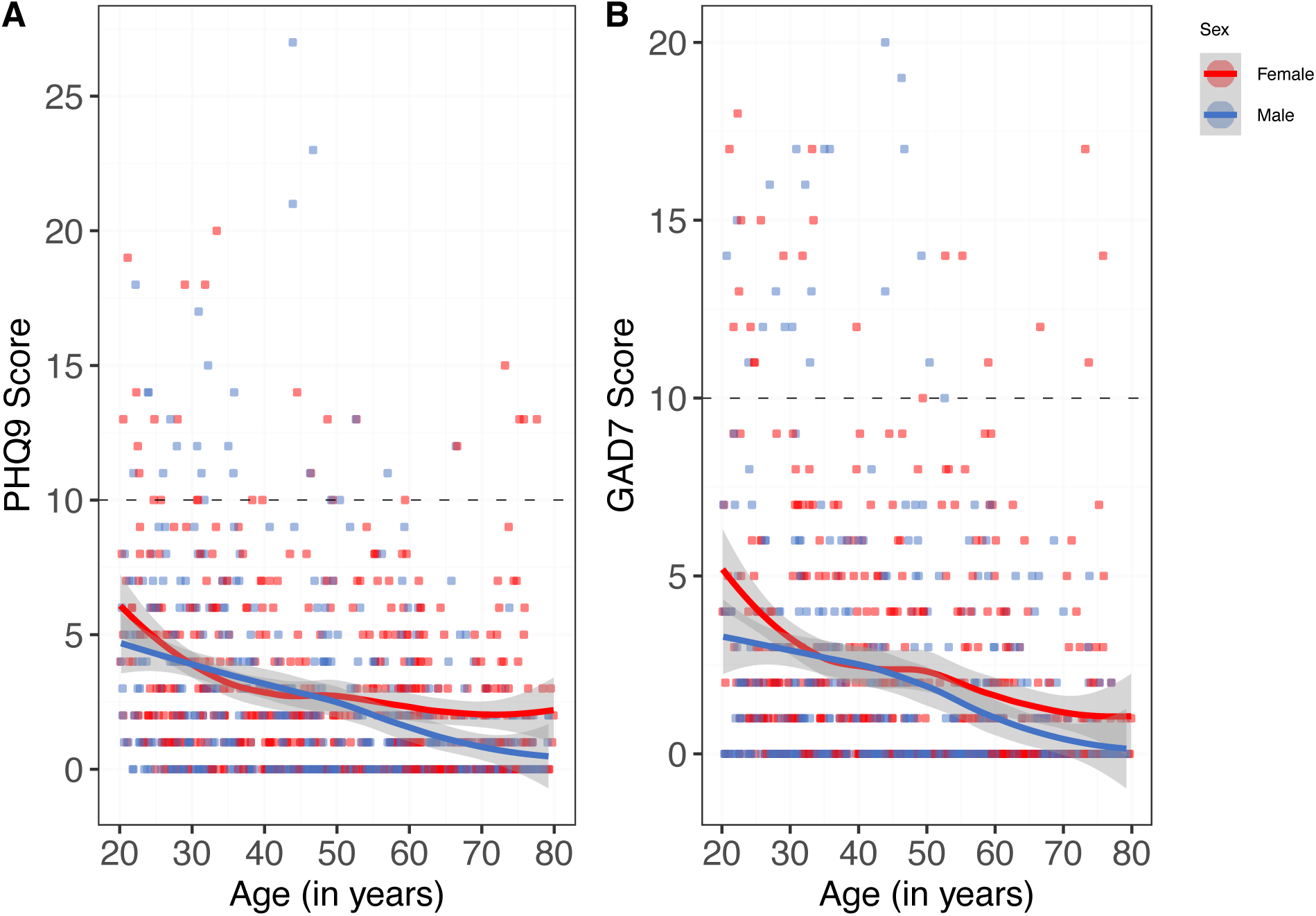
Anxiety and depression scores of HHGP-HK donors. (A) GAD7 and (B) PHQ-9 scores as a function of age and sex in HHGP-HK donors. (n=1025)

**Table S1.** Results of the pilot study.

| Call Centre | Count | Research Clinic | Count |
| --- | --- | --- | --- |
| Complete interview (I) | 8 | V0 completed | 8 |
| Partial interview (P) | 0 | V1 completed | 4 |
| Refusal and break-off (R) | 41 | V1 pending | 4 |
| Non-contact (NC) | 38 |  |  |
| Other (O) | 7 |  |  |
| Ineligible | 7 |  |  |
| Total | 101 | V1 total | 8 |
| **Response rate (RR)** | **8.51%** | **Enrolment rate (ER)** | **8.51%** |
| **Cooperation rate (CR)** | **16.3%** | **Inclusion rate (IR)** | **100%** |

**Table S2.**
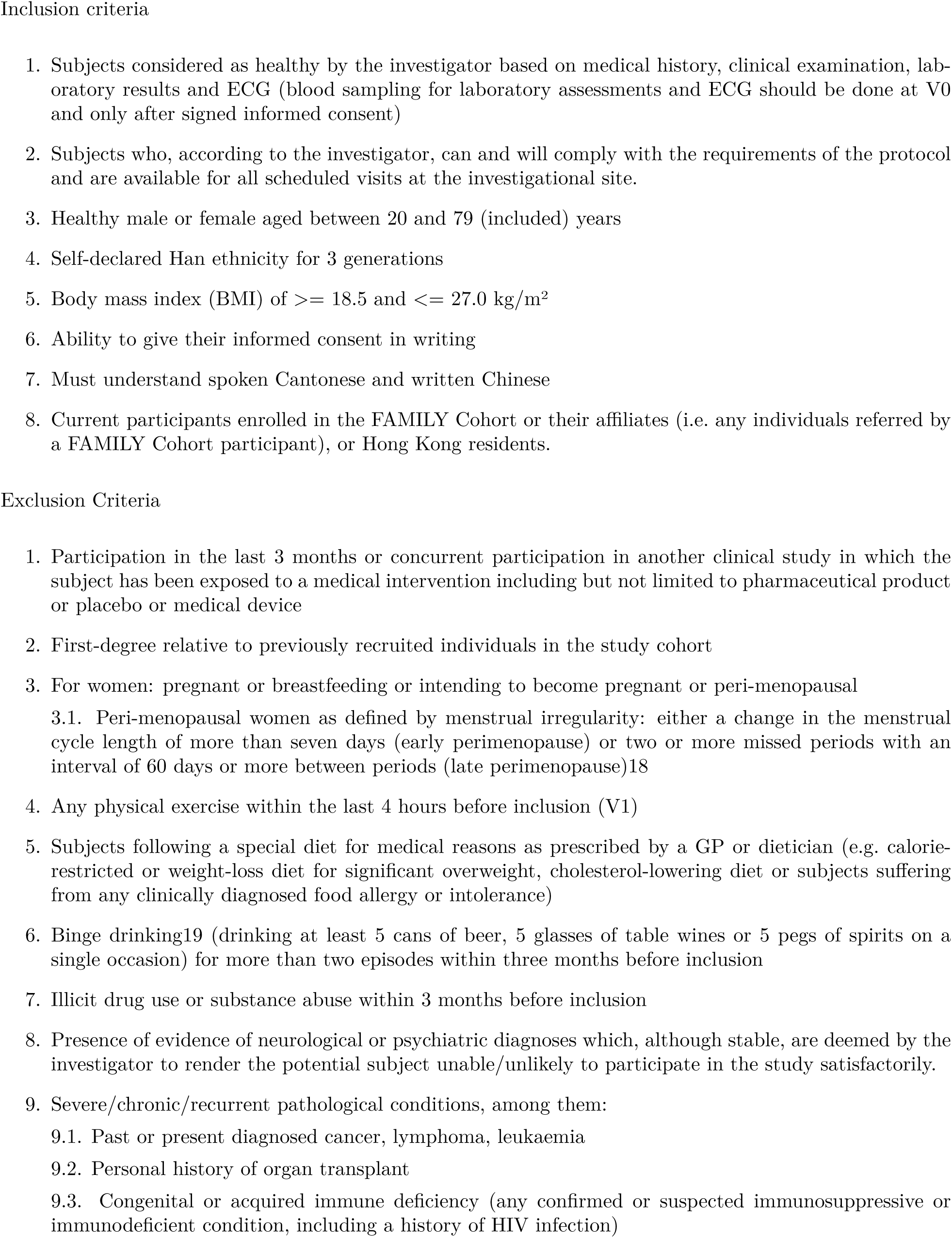
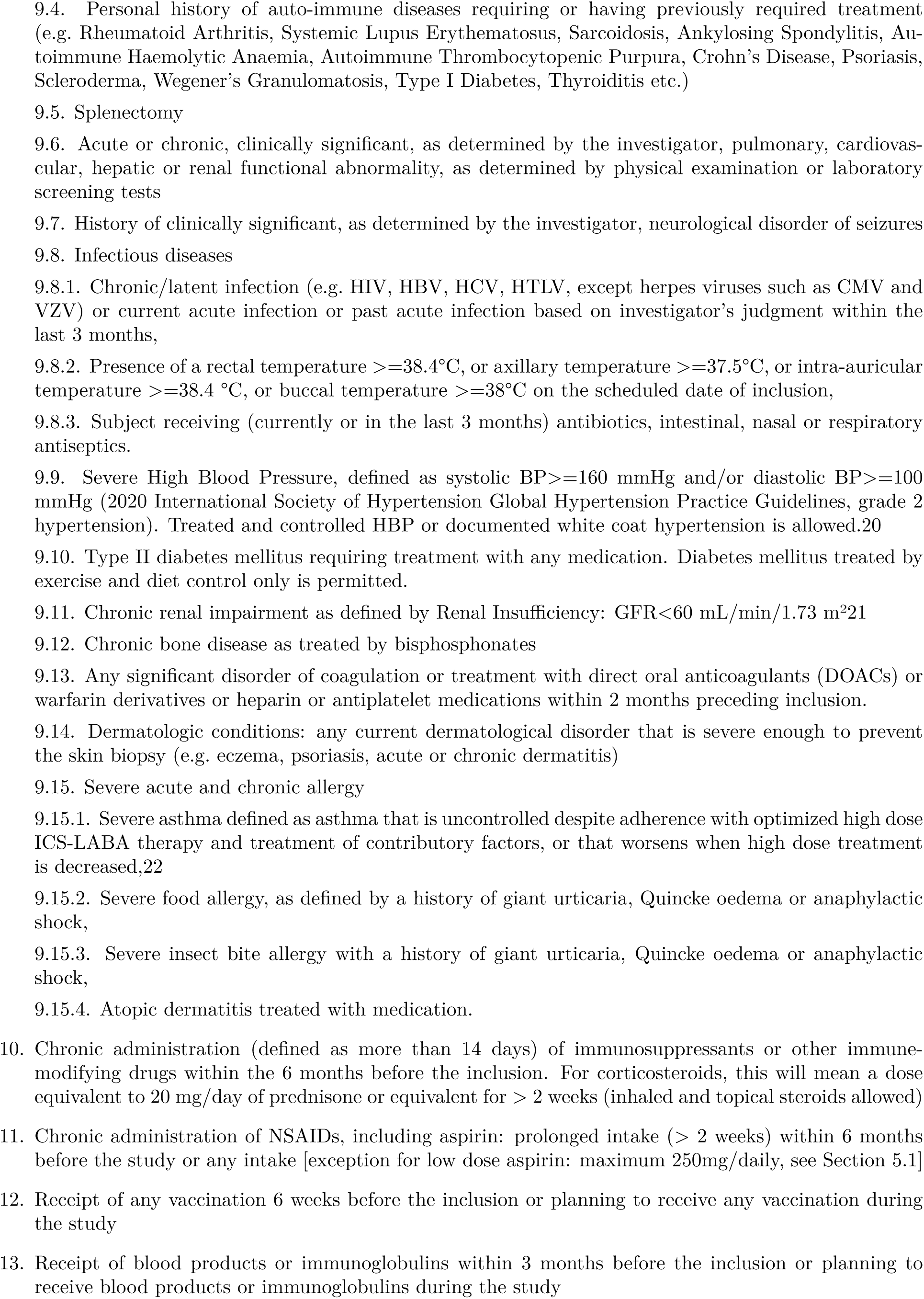
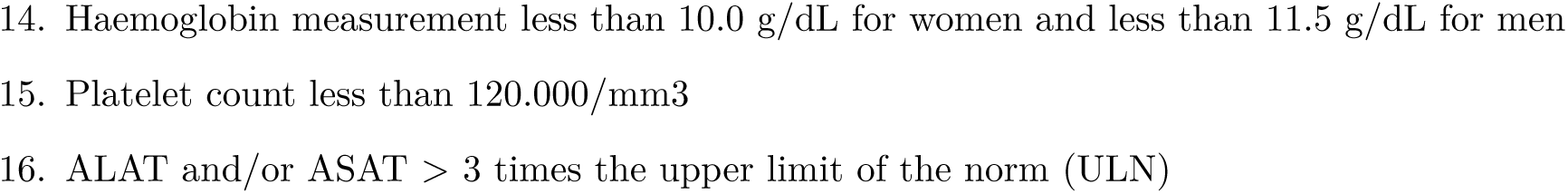
Complete inclusion and exclusion criteria of the HHGP-HK study.

**Table S3.** Reasons for V1 enrolment attrition (148) and withdrawal (22)

| **Cause** | **Count** | **Remarks** |
| --- | --- | --- |
| BMI out of range (Target 18.5 - 27.0) | 28 | 17x >upper limit (30.3, 29.1, 28.8, 28.8, 28.8, 28.7, 28.7, 28.1, 28.0, 27.9, 27.9, 27.8, 27.8, 27.5, 27.4, 27.2, 27.2); 11x < lower limit (18.2, 18.0, 18.0, 17.9, 17.8, 17.7, 17.6, 17.5, 17.4, 17.2, 17.0) |
| Recent infection/ unmatched recruitment timeline | 26 | 16x COVID-19; 4x recent infection requiring antibiotic use |
| Communication contact failure/ loss to follow up | 23 | 11x not completed V0; 11x completed V0; 1x no show for 3 times or more |
| Withdrawal request/ no longer willing to participate | 22 | 8x too busy; 2x too much blood taking/ complicated procedures; 2x difficult phlebotomy; 2x no DXA offered; 2x undisclosed personal reasons; 1x lack of confidence; 1x family not allowed; 1x recent pregnancy; 1x expectation different in results reporting; 1x losing confidence due to V0 specimen clotting; 1x inconvenient timing from recent surgery |
| Autoimmune diseases requiring likely treatment as revealed during V0/V1 consultation or through blood investigations | 15 | 12x thyroid dysfunction; 1x newly diagnosed ulcerative colitis; 1x rheumatoid arthritis; 1x nonspecific autoimmune condition on steroid treatment |
| Clinically significant pathological condition(s) or history of cancer as revealed during V0/V1 consultation or through blood investigations | 13 | 1x hypercalcemia with increase in ALP; 1x history of bladder cancer; 1x suspected psoriasis flare; 1x eczema flare up; 1x uncontrolled blood pressure; 1x suspected uveitis; 1x deranged liver function test; 3x renal insufficiency; 1x post stroke seizure; 1x bisphosphates treatment; 1x lung cancer |
| Pancytopenia/ bicytopenia or thrombocytopenia | 10 | Platelet count less than 120,000/mm<sup>3</sup> for thrombocytopenia |
| Low haemoglobin | 8 | <10.0g/dL for women and <11.5g/dL for men |
| Chronic infection | 10 | 9x HBsAg positive; 1x HIV-1 & 2 Ab/ p24 positive |
| Abnormal ECG | 5 | 1x Ischemic changes/ congenital heart condition; 3x newly diagnosed atrial fibrillation; 1x hypertensive left ventricular hypertrophy |
| History of anaphylaxis/ severe allergic reactions to food | 3 | No further observations |
| Miscellaneous | 10 | 3x 1<sup>st</sup> degree relative enrolment; 2x perimenopausal; 2x binge drinking; 2x voided blood samples from site power failure; 1x diabetic medication |

**Table S4.** Abbreviations of clinical laboratory measures.

| Abbreviation | Name | unit |
| --- | --- | --- |
| ALB | Albumin | g/L |
| ALP | Alkaline Phosphatase | U/L |
| ALT | Alanine Aminotransferase | U/L |
| AST | Aspartate Aminotransferase | U/L |
| BASO COUNT | Basophils Count | $10^9/L$ |
| BASO PERC | Basophils Percentage | % |
| BICARB | Bicarbonate | mmol/L |
| BILI | Total Bilirubin | umol/L |
| BILI DIRECT | Direct Bilirubin | umol/L |
| BMI | Body Mass Index | kg/m<sup>2</sup> |
| BUN | Blood Urea Nitrogen | mmol/L |
| C2i | Centre for Immunology & Infection | N/A |
| CA | Calcium | mmol/L |
| CBC | Complete Blood Count | N/A |
| CHOL | Total Cholesterol | mmol/L |
| CHOL HDL Ratio | Total Cholesterol/High-Density Lipoprotein Ratio | N/A |
| CL | Chloride | mmol/L |
| CMV Ab | Cytomegalovirus IgG Antibodies | N/A |
| CNIL | Commission Nationale de l'Informatique et des Libertés | N/A |
| CREAT | Creatinine | umol/L |
| CRF | Case Report Form | N/A |
| CRP | C-Reactive Protein | mg/L |
| CSE | Centre for Sports and Exercise | N/A |
| CVD | Cardiovascular Disease | N/A |
| DXA | Dual-energy X-ray Absorptiometry | N/A |
| ECG | Electrocardiogram | N/A |
| EDTA | Ethylenediaminetetraacetic Acid | N/A |
| EOS COUNT | Eosinophils Count | $10^9/L$ |
| EOS PERC | Eosinophils Percentage | % |
| FDR | False Discovery Rate | N/A |
| FG500 | Human Functional Genomics Project | N/A |
| GAD-7 | General Anxiety Disorder-7 | N/A |
| GGT | Gamma-Glutamyl Transferase | U/L |
| Globulin | Globulin | g/L |
| GLUC | Fasting Plasma Glucose | mmol/L |
| Haematocrit | Haematocrit | % |
| Haemoglobin | Haemoglobin | g/dL |
| HDL | High Density Lipoprotein | mmol/L |
| HHGP-HK | Human Global Project - Hong Kong | N/A |
| HBV | Hepatitis B | N/A |
| HCV | Hepatitis C | N/A |
| HIV | Human Immunodeficiency Virus | N/A |
| HTLV I/II | Human T-Lymphotropic Virus I & II | N/A |
| HKU/HA HKW IRB | Institutional Review Board of The University of Hong Kong/Hospital Authority Hong Kong West Cluster | N/A |
| HKUCTR | Clinical Trials Centre of the University of Hong Kong | N/A |
| HRV | Heart Rate Variability | ms |
| IgA | Immunoglobulin A | g/L |
| IgG | Immunoglobulin G | g/L |
| IgM | Immunoglobulin M | g/L |
| K | Potassium | mmol/L |
| LDL | Low Density Lipoprotein | mmol/L |
| LYMPHO COUNT | Lymphocytes Count | 10<sup>9</sup>/L |
| LYMPHO PERC | Lymphocytes Percentage | % |
| MCH | Mean Corpuscular Haemoglobin | pg |
| MCHC | Mean Corpuscular Haemoglobin Concentration | g/dL |
| MCV | Mean Cell Volume | fL |
| MID | Monocytes Percentage | % |
| MONO COUNT | Monocytes Count | 10<sup>9</sup>/L |
| MPV | Mean Platelet Volume | fL |
| NEUTRO COUNT | Neutrophils Count | 10<sup>9</sup>/L |
| NEUTRO PERC | Neutrophils Percentage | % |
| PBMC | Peripheral Blood Mononuclear Cell | N/A |
| PCA | Principal Component Analysis | N/A |
| PCR | Polymerase Chain Reaction | N/A |
| PHOS | Phosphorus | mmol/L |
| PHQ-9 | Patient Health Questionnaire-9 | N/A |
| PLAT | Platelet Count | $10^9/L$ |
| PPE | Personal Protective Equipment | N/A |
| PROT | Total Protein | g/L |
| R/LFT | Renal/Liver Function Test | N/A |
| RBC | Red Blood Cell Count | $10^{12}/L$ |
| RDW | Red Blood Cell Distribution Width | % |
| RMSSD | Root Mean Square of Successive Differences | ms |
| SNP | Single Nucleotide Polymorphism | N/A |
| SODIUM | Sodium | mmol/L |
| Total IgE | Total Immunoglobulin E | IU/mL |
| TRIG | Triglycerides | mmol/L |
| TSH | Thyroid Stimulating Hormone | mIU/L |
| URATE | Urate | umol/L |
| UREA | Urea | mmol/L |
| V0 | Enrolment Visit 0 | N/A |
| V1 | Inclusion Visit 1 | N/A |
| VLDL | Very-Low-Density Lipoprotein | mmol/L |
| WBC | White Blood Cell Count | $10^9/L$ |

**Supplementary appendix:** case report form

## Notes

### Competing Interest Statement

The authors have declared no competing interest.

### Author Declarations

The HHGP-HK clinical study protocol was first approved by the Institutional Review Board of The University of Hong Kong/Hospital Authority Hong Kong West Cluster (HKU/HA HKW IRB) on 16 August 2021 with reference number UW21-549. HHGP-HK is registered with the HKU Clinical Trial Registry (HKUCTR) and is available for public access on www.HKUCTR.com with study identifier HKUCTR-2959. The study is sponsored by the Centre for Immunology & Infection (C2i) and was conducted as a single center study at the University of Hong Kong without any investigational product. The protocol is registered under ClinicalTrials.gov (study number NCT05174624). Three subsequent HKU/HA HKW IRB clinical research ethics review approval amendments were obtained to adapt to recruitment challenges due to the COVID-19 pandemic.

